# COVID-19 mortality according to civilian records

**DOI:** 10.1101/2020.08.07.20170183

**Authors:** Lisandro Lovisolo, Diego H. S. Catalão, Rodrigo B. Burgos, Malú Grave, Pamella Constantino-Teles, Americo Cunha

## Abstract

In this short report, we bring some data-driven analyses of COVID-19 mortality in Brazil. The impact of COVID-19 is evaluated by comparing the 2019 and 2020 civilian death records. There is evidence of a considerable excess of deaths since the pandemic started with respect to the previous year. In some states, it is clear that not all excess of deaths in 2020 is due to COVID-19, but to other respiratory causes that did not present the same prevalence in the previous year. Because of this unusual behavior of respiratory deaths, we may infer the evidence of a huge amount of under-reporting deaths due to the COVID-19. The data also shows that COVID-19 has produced an excess death in all ages besides people above 90 and below 10 years. In addition, when separates by sex, data indicate a larger increase in the deaths among males than females.

## Introduction

The spread of the SARS-CoV-2^1,2^ around the world occurred rapidly^3^ and on a planetary scale not seen since the Influenza pandemic (the so-called *Spanish flu)* between 1918 and 1920, such that on 3 March 2020, the World Health Organization (WHO) decreed a pandemic state^4^ concerning the associated respiratory disease, named COVID-19^5^. This fact induced significant changes in the social, economic, and cultural order, once practically all the countries were forced to adopt non-pharmacological interventions (NPI)^6^ to avoid a possible collapse of their respective health systems, and to prevail (or at least mitigate) a massive loss of human life^7^.

In Brazil, the first official notification occurred at São Paulo municipality on 25 February 2020, when a 61 years-old man tested positive for the infection of SARS-CoV-2^8^. The first death was recorded 21 days later^9^, on 17 March 2020, when the country registered 293 official cases. Since then, the SARS-CoV-2 has spread to all Brazilian states^10–12^, achieving around 3 million reported cases and 100 thousand deaths by the time of submission^13,14^. These numbers that make Brazil the second nation in the numbers of officially reported cases and deaths by early August 2020.

The need to know the global and localized impact of the disease urged means to monitor the progress of the pandemic. In this sense, health authorities around the world monitored various indicators of the pandemic progress in different areas. The numbers of cases and deaths are the most used to show the current epidemic status in a region of interest and the level of severity that this region is affected by the COVID-19 disease^13,15^. Although the actual numbers may differ from the reported ones, due to under-reporting, they have been the most common tool to track the spread of the COVID-19 pandemic. These have been used purely, in normalized forms, and to compute other indices as the reproduction number^16^ to track the spread of COVID-19.

Among the indices being employed to track the spread of COVID-19 pandemic, the number of deaths seems to be the most reliable for evaluating the actual effects of the COVID-19^17^. The report of deaths is compulsory almost everywhere, while the report of infections may rely on the disease severity, extension of population testing, and other aspects. The number of deaths is also essential to estimate epidemiological quantities such as mortality and lethally, which help to map the severity of the disease. These aspects make the number of deaths good to track the COVID-19 impacts on a country or a region^18^. Nevertheless, different countries have adopted different procedures for accounting deaths as resulting from SARS-CoV-2^19^. While at some, any suspicious death is reported as due to the COVID-19 epidemic, others require testing to track it precisely. In large area countries, like Brazil, the protocol may also change across states and cities. Still, this number is less susceptible to the inevitable under-reporting than the number of cases.

In this short report, we bring some analyses on COVID-19 data in Brazil. More specifically, we monitor the Brazilian National Civil Record, an open-source data of civilian life records, named the *Portal da Transparência* - *Registro Civil*^20^. It aggregates the registration of births, marriages, and deaths. We query it for the number of deaths reported between January and June for the years 2019 and 2020 in the different states of Brazil. The obtained data are employed to evaluate the impact of COVID-19 for male and female and different age intervals. It also serves as evidence for the vast under-reporting, since, in some states, it is clear that not all excesses deaths (the difference between the number of deaths and the expected from the previous year) are due to COVID-19, but instead to other respiratory causes that did not present the same prevalence in the previous year, showing abnormal behavior.

## Methods

In this work, we compare the number of daily deaths reported in Brazil between January and June of 2020, under the influence of the COVID-19 epidemic, with the number of deaths reported in Brazil over the same period of 2019. For such a task, the following methodology is employed. We present a comparison between the numbers in 2019 and in 2020 since the data available in the *Portal da Transparência* - *Registro Civil*^20^ presents inconsistencies in previous years as it has become operational in 2018.

### Civilian records

To evaluate the impacts on different groups, we try to explore at most the 2019 and 2020 data available in the *Portal da Transparência* - *Registro Civil*^20^. It would be interesting to make this comparison with the other years of the last decade, but these records are not available in this database. It is worth mentioning that due to legal deadlines, there is a significant delay between a death occurrence and its report, which may take more than one month in some cases. After someone dies, his/her family has to notify the notary’s office within 24 hours after the occurrence. However, this deadline may be extended to 15 days (or even 30 days in some particular cases). The notary’s office has five days to register the death in the records, and eight more days may be needed for the registry to enter the online National Civil Record. Due to the limitations imposed by the pandemic, naturally, these deadlines have been extended. Therefore, we know that even considering data acquired in July, the deaths that occurred in May may not have reached their final number. Although such a delay occurs (as one observes from the decay in the number of deaths for 2020 in the graph correspondent to Minas Gerais (MG) in Figure 2), the considered data still provides a reasonable way to track the evolution of the number of deaths from SARS-CoV-2 in Brazil.

### Data extraction

The data are collected by employing a simple boot that queries the *Portal da Transparência* - *Registro Civil*^20^ for the deaths on a given date. This procedure is done for all causes in all Brazilian Federation states. Another boot does the same process to query this information for sexes and ages.

### Data analyses

We analyze the number of deaths during the pandemic according to the following categories:

1. *Total number of deaths:* considers all the deaths that have occurred in a given time interval;
2. *Aggregate number of deaths from respiratory causes:* it aggregates deaths due to pneumonia, SARI (Severe Acute Respiratory Infection) and respiratory insufficiency for 2019, and it also includes COVID-19 for 2020;
3. *Number of deaths from respiratory causes:* in addition to the aggregate number of deaths from respiratory causes, we evaluate their changes separately from 2019 to 2020. This is done for the four aforementioned respiratory causes for death;
4. *Control groups:* to access the database consistency and validity, we analyze the changes in the number of deaths due to unknown causes, sepsis, and all other reasons.

## Data processing

The present analyses split the data into female and male sexes and also into groups by age. For the last, we employ the intervals of 0–9, 10–19, 20–29, 30–39, 40–49, 50–59, 60–69, 70–79, 80–89, 90–99, ≥ 100 and N.A. (when the age is not available). For each group, we compare the changes in the absolute and relative number of deaths for different causes between 2019 and 2020. From these changes one can observe the effects of COVID-19 among the different groups.

Due to Brazil’s large area, the disease has arrived at different dates in different regions^11,21^. Moreover, the heterogeneity of Brazil reveals different paces of the epidemic in the several states of the country and regions within states, so that the COVID-19 epidemic is composed of different surges in different regions, states, and even municipalities within the Brazilian territory. Therefore, we analyze the data of the whole country and the states separately.

Cross-correlation is employed to evaluate the statistical dependence between percentage variations in the number of deaths due to different causes. Let the change in the total number of deaths in a given state and month in 2020 w.r.t 2019 be denoted by Δ_tot_, _month_(state) and that the change in the number of deaths for another cause in a given state and month in 2020 w.r.t 2019 be denoted by Δ _cause_, _month_ (state). The cross correlation between both is computed using

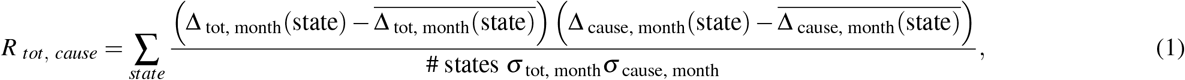

where the summation occurs over the states, the over-line (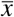) denotes the average operator (or expectancy), and # states is the number of states (27 in Brazil). Above, *σ*_tot, month_ is the standard deviation of Δ_tot, month_(state), i.e.,

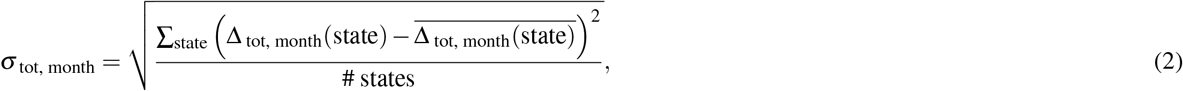

and *σ*_cause, month_ (state) is defined accordingly.

## Results and discussion

### The inception of COVID-19 in Brazil

The number of daily deaths in Brazil, from January to June of 2019 and 2020, is shown in Figure 1. At the top of this Figure, around the last week of March 2020, one readily sees that the 2020’s total daily deaths curve departs from its 2019’s counterpart. This effect is highly pronounced in the trace of daily deaths from respiratory causes. As one can see at the bottom graph in the same Figure, the main cause of deaths in the last set is due to SARS-CoV-2. Nevertheless, there are two other important aspects to observe. First, the number of daily deaths reported from pneumonia has an increase during the mid-March 2020 but decreases as the number of deaths attributed to COVID-19 increases. This decrease gets more pronounced as April 2020 starts, and the number of pneumonia daily deaths goes below the one observed in the previous year. Second, SARI appears as responsible for several daily deaths during 2020, while this was not observed in the previous year. These two observations suggest that, in the first two weeks of the Brazilian epidemic, cases of COVID-19 may have been wrongly reported as pneumonia, and the opposite occurring shortly after. These data suggest that, even with the epidemic already in evidence in the news, part of the Brazilian health system still took a few weeks to be able to differentiate COVID-19 from other respiratory diseases.

**Figure 1.**
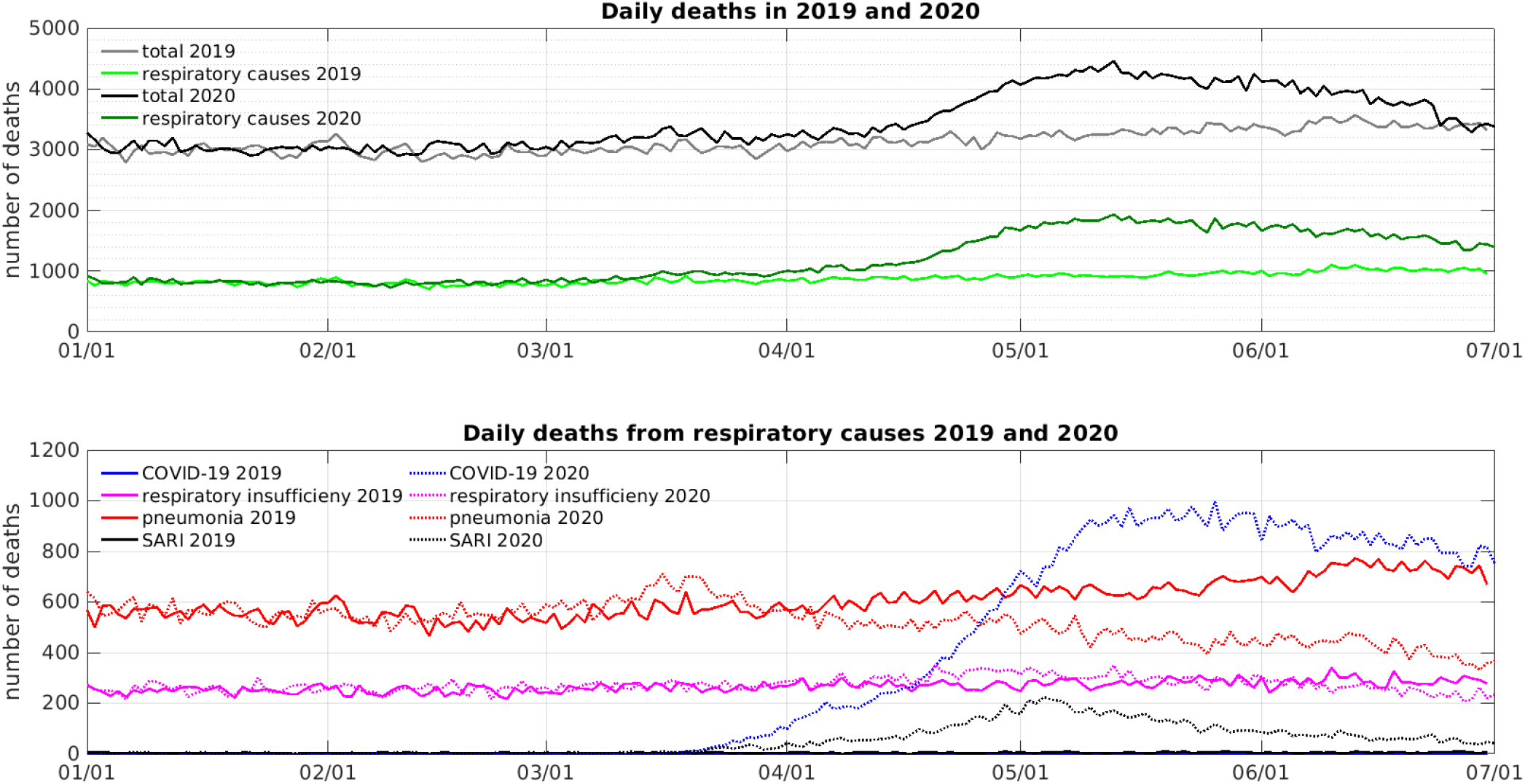
Daily deaths in Brazil between January and June of 2019 and 2020. At the top, we present the daily deaths from all causes, and the aggregate number of daily deaths due to respiratory causes (respiratory insufficiency, pneumonia, SARI, and COVID-19). At the bottom, we present the daily deaths due to respiratory causes (respiratory insufficiency, pneumonia, SARI, and COVID-19) separately. There is clearly an increase in total mortality caused by an increase in deaths from respiratory causes. Data gathered on July 30th, 2020 from^20^.

The data presented in the top graph in Figure 1 show that the COVID-19 epidemic makes the number of daily deaths in Brazil to increase in almost one third. Moreover, they show that this increase is essentially due to respiratory causes. Complementary, the data producing the bottom graph in Figure 1 suggests that the increase in daily deaths is highly correlated with the disruption of the COVID-19 epidemic in the different Brazilian states.

### Spread of the disease across the country

In each Brazilian state, the COVID-19 has arrived at a different time, producing different effects, as shown in Figure 2, which presents the evolution of daily deaths in the Brazilian states between January and June of 2019 and 2020. The analysis described in the last section can be held for every state.

**Figure 2.**
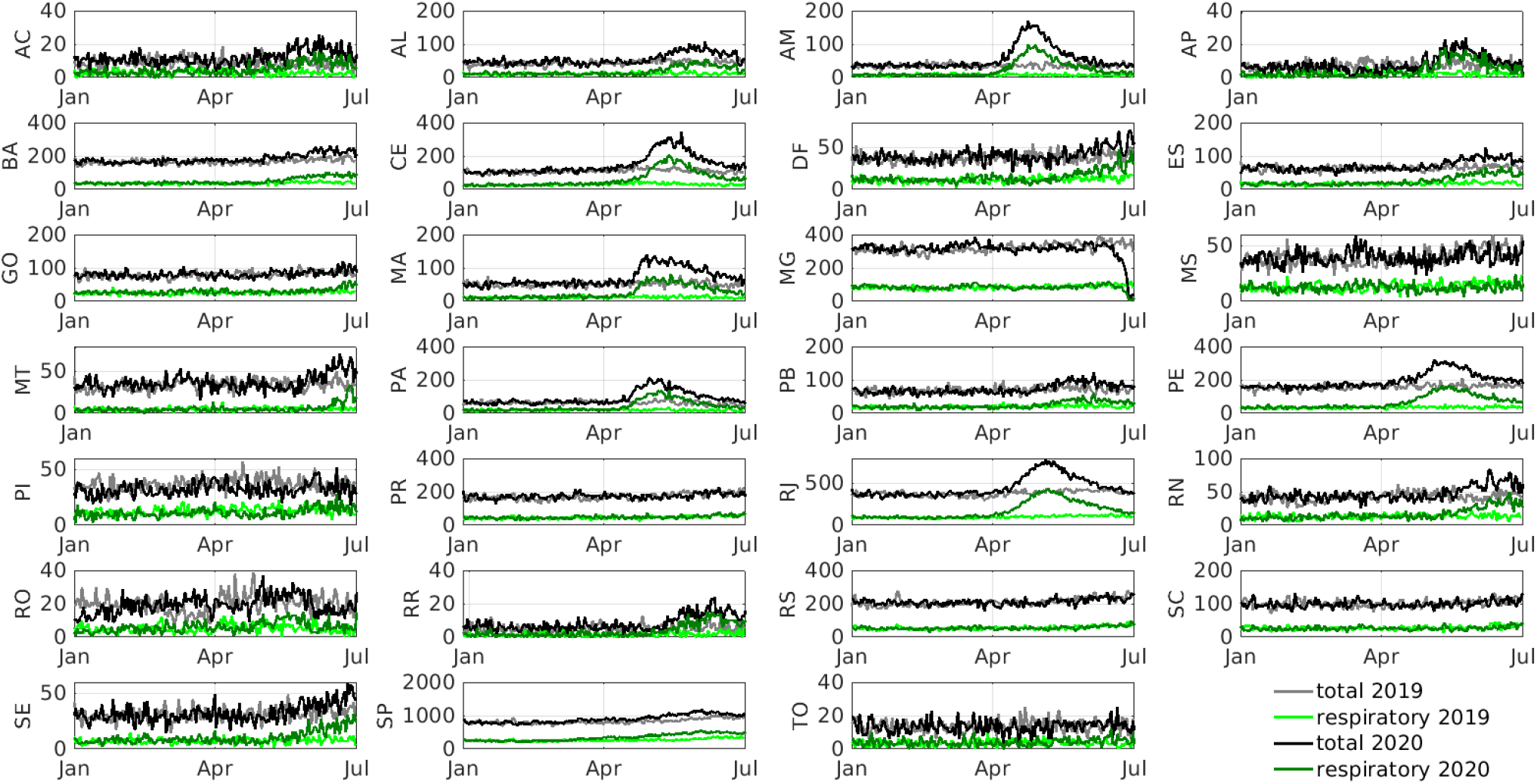
Daily deaths in the Brazilian states between January and June of 2019 and 2020. Each graph shows the number of daily deaths from all causes, and the aggregate number of daily deaths due to respiratory causes (respiratory insufficiency, pneumonia, SARI, and COVID-19). The vertical labels show the state abbreviations. Data gathered on July 30th, 2020 from^20^.

Tables 1 and 2 in the supplementary material present such analyses of the number of deaths due to different causes in the months April, May, and June, comparing 2020 with 2019 grouping the states within Brazilian macro-regions. While in Table 1 (supplementary material), one can access the numbers of total deaths and respiratory cause deaths, in Table 2 (supplementary material), the numbers are desegregated among the different respiratory causes considered.

### Impact in the monthly number of deaths

Figure 3 shows the percentage change in the total number of deaths and in the number of deaths due to respiratory causes for the 27 Brazilian states in April, May and June 2020 w.r.t to 2019 (the raw numbers provided in Table 1 in the supplementary material). The maps in the top row of Figure 3 present the percentage change in the total number of deaths, and the maps in the bottom row present the percentage increase in the number of deaths due to respiratory causes. Clearly, the percentage increase in the number of deaths due to respiratory causes is greater than the one in the total number of deaths, since the first is a subset of the last. However, the graphs suggest a correlation between the change in total mortality and respiratory causes. To investigate that, we evaluate the cross-correlation between the variation in the total number of deaths and the variation in the number of deaths due to respiratory causes in 2020. In April, the cross-correlation is 0.9503, in May, it is 0.9447, and in June, 0.8935. These cross-correlation values are very high and show that the increase in the total number of deaths is greatly influenced by the increase in the number of deaths from respiratory causes.

**Figure 3.**
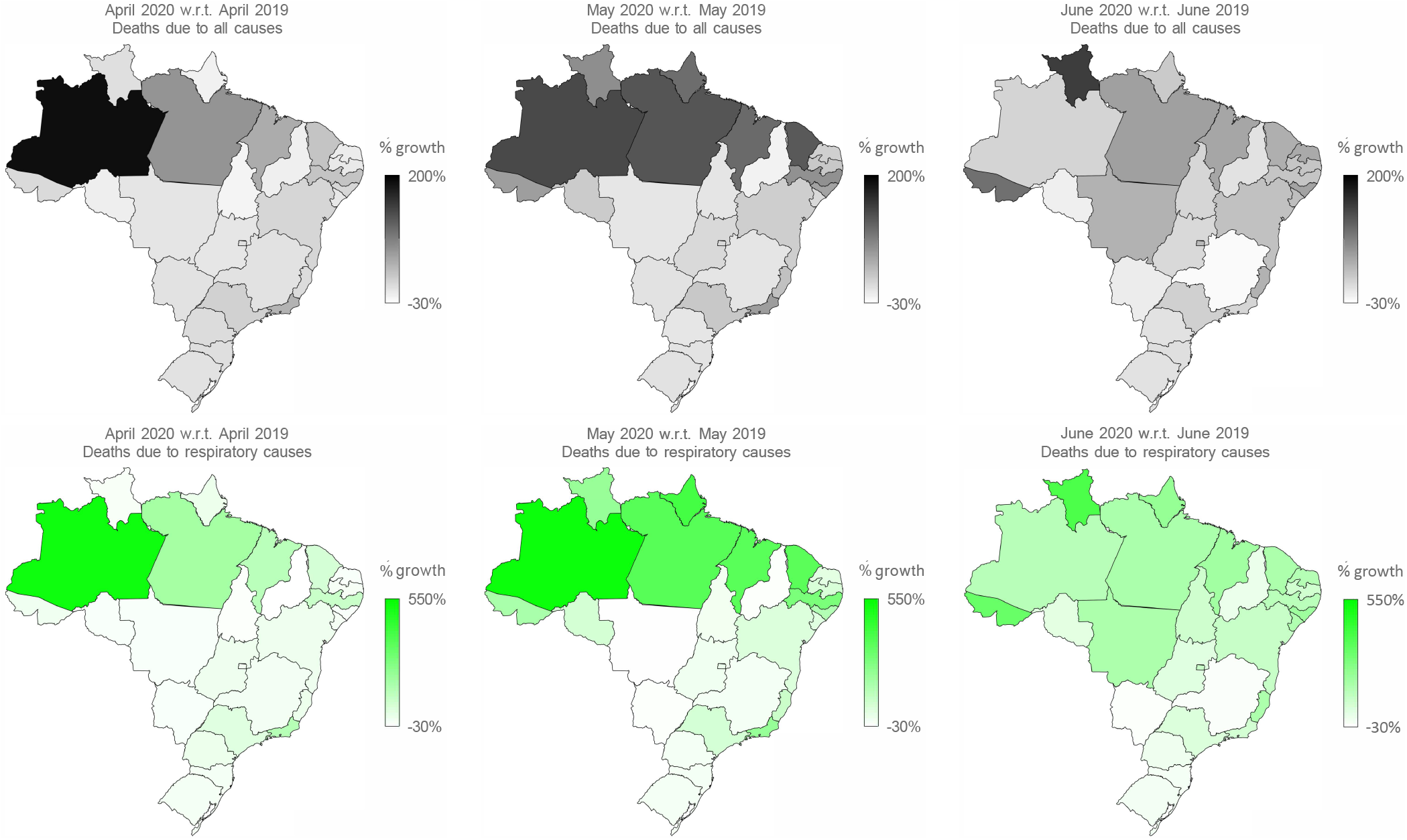
Monthly increases in the total number of deaths and the number of deaths due to respiratory causes in the Brazilian states in April, May and June of 2020 w.r.t 2020. The maps in the top row present the percentage change in the total number of deaths, and the maps in the bottom row present the percentage increase in the number of deaths due to respiratory causes. Data gathered on July 30th, 2020 from^20^.

To confirm if the previous high cross-correlation values are relevant, we also compute cross-correlation between the variation in the total number of deaths in one month and variation in the number of respiratory deaths in another month, that is considering a time lag between the series. The value of the cross-correlation when considering the number of deaths of April and May is 0.6970; when considering the number of deaths of April and June, it is 0.1283; May and April, 0.6539; May and June, 0.5675; June and April, −0.0201; and, at last, June and May, 0.3199. One readily notices that these cross-correlation values are reasonably smaller than the ones computed within the same month and that they decrease as the lag between the months increases. The smaller correlations values obtained when considering different months allow us to infer that deaths due to respiratory causes are the main source of the increase in total deaths.

Furthermore, we evaluate the cross-correlation of the variation in the total number of deaths and the variation in the number of deaths due to “other” causes and sepsis. It shall be noticed that “other” causes is the main contributor for the number of deaths since it consider all causes not included in the respiratory group, neither sepsis, since the last is a consistent cause for deaths over time; therefore considering both in the analyses provides ways to detect changes in mortality patterns. In April, the first cross-correlation (variation in the total number of deaths and the number of deaths from other causes) is 0.9209; in May it is 0.7066; and in June, 0.7069. For the second cross-correlation (variation in the total number of deaths and the number of deaths from sepsis), one obtains 0.1776 in April, −0.2045 in May, and 0.0675 in June, showing that there is no apparent link between these causes of deaths.

On one hand we observe that a reduction in the correlation between the change in the total number of deaths and the change in the number of deaths due to other causes is a clear indication of the change in mortality provoked by the COVID-19. On the other hand, we observe a great correlation between the change in the total number of deaths and the change in the number of deaths due to respiratory causes. Moreover, the last does not hold when the lags between months is one or two. These results indicate that the observed increase in the total number of deaths in each Brazilian state is highly correlated with the increase in the number of deaths from respiratory causes that is boosted by the inception of the COVID-19 in each state.

### Deaths by age intervals

Figure 4 presents the number of deaths in Brazil between January and June of 2019 and 2020, separated by sex and among ranged ages. The graphs in Figure 4 show the number of deaths reported as being due to respiratory causes (respiratory insufficiency, pneumonia, SARI, and COVID-19, the last only for 2020), sepsis, others (all but the previously mentioned causes) and from unknown causes. A small change can be seen in the age groups below 30 years. On the other hand, in all other age groups, we can see a pronounced increase in mortality, with more significant expansion in the age groups above 60 years, where the COVID-19 is the leading cause of deaths.

**Figure 4.**
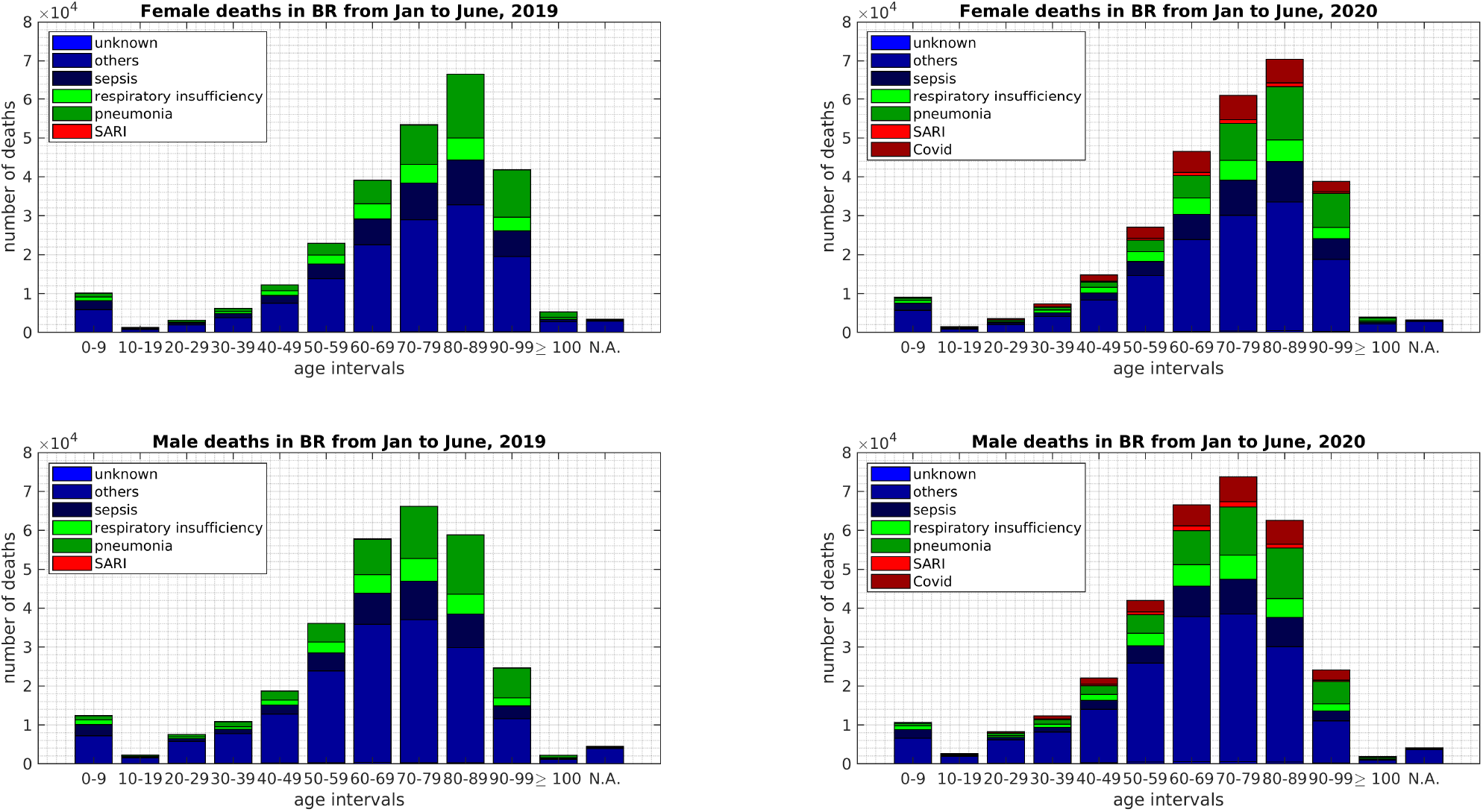
Number of deaths from different causes in Brazil between January and June of 2019 and 2020, for both sex and ranged by age intervals of 10 years. At the top left corner, we present the number of female deaths resulting from different causes in 2019 (see methods). At the top right corner, we present the number of female deaths resulting from different causes in 2020 (including COVID-19). At the bottom left corner, we present the number of male deaths resulting from different causes in 2019. At the bottom right corner, we present the number of male deaths resulting from different causes in 2020 (including COVID-19). Data gathered on July 30th, 2020 from^20^.

Figure 5 provides another view of the data in Figure 4, as it compares the total number of deaths against the number of deaths due to respiratory causes, and due to COVID-19. The number of deaths in almost all age intervals increases in 2020 w.r.t 2019, except in the extreme ranges (0–9, 90–99 and 100–109). This is readily shown in the graphs at the top and in the middle of Figure 5. The middle graph presents the changes in the number of deaths between 2019 and 2020 from all causes, respiratory only, and COVID-19, for each sex and age interval. For all age intervals, we may observe an increase in the number of deaths in 2020. We also see that the increase is principally due to respiratory causes and, among them, mostly is due to COVID-19. The change in the number of deaths is more significant for the age ranging from 60 to 69 years, followed by the range 70–79, then by 80–89 (female) and 50–59 (male), and 50–59 (female) and 80–89 (male), and then in the age intervals 40–49, 30–39 and 20–29. Nevertheless, surprisingly, although there are many deaths due to COVID–19 in the age range from 90 to 99 years old, the net effect in this age range is negative, i.e., fewer people with 90 to 99 years have died in 2020, during the COVID–19 outbreak in Brazil, than in 2019.

**Figure 5.**
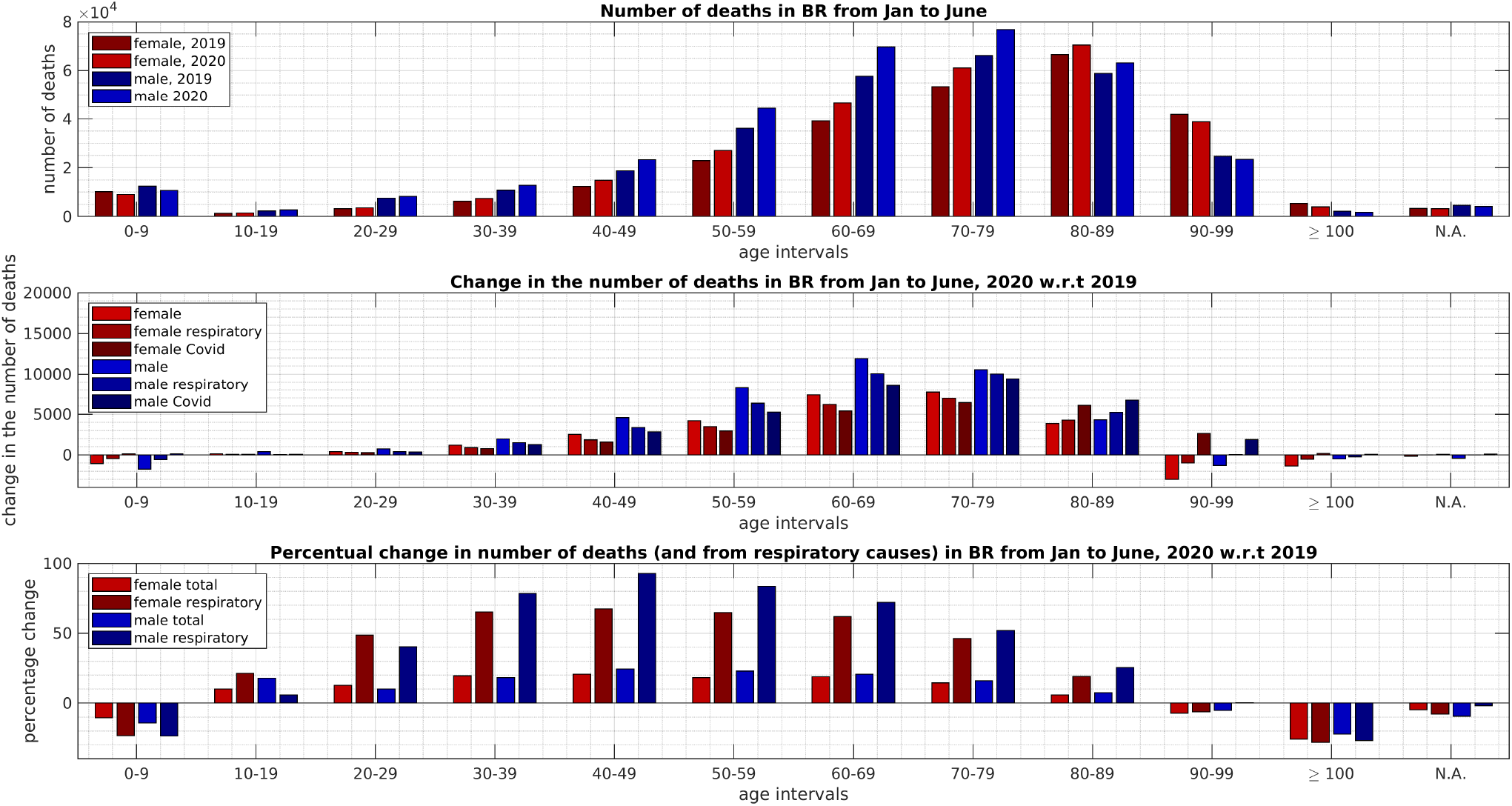
Deaths in Brazil between January and June of 2019 and 2020 for both sexes ranged by intervals of 10 years. At the top, we present the total number of deaths for both sexes. At the middle, we present the changes in the number of deaths between 2019 and 2020 from all causes, respiratory only, and COVID-19. At the bottom, we present the percentage change within each group (age and sex) in the total number of deaths and in the number of deaths reported as being due to respiratory causes. Data gathered on July 30th, 2020 from^20^.

Meanwhile, the graph at the bottom in Figure 5 shows that the percentage change in the number of deaths is above 10% in all adult age intervals, i.e., 20–29, 30–39, 40–49, 50–59, 60–69 and 70–79. In all of these, the percentage change in the deaths due to respiratory causes (which includes COVID-19) is even more significant, representing its main constituent as one apprehends from the middle graph. At last, it is essential to emphasize that the increase in deaths due to respiratory causes is more pronounced in the ages ranging from 30 to 69 years old, where it lies above 50% for both sexes. For the 20–29 years old range, deaths by respiratory causes have also increased significantly, above 40% for both sexes, producing a percentage increase in the total deaths larger than 10% for both sexes. This change is larger than the expected by natural demographic reasons^22^.

### Deaths by respiratory causes

The graphs in Figure 6 result from the number of deaths attributed to the different respiratory causes in Brazil in 2019 and 2020. One notices that there is a small decrease in the number of deaths attributed to pneumonia from 2019 to 2020; the higher the age, the greater is this decrease for both sexes, but a change occurs for females above 100 years and males above 90 years. However, in 2020 there is a large number of deaths attributed to SARI (Severe Acute Respiratory Infection), which are not observable in 2019 for both sexes. Meanwhile, there is an even larger number of deaths due to COVID-19. Except for the age intervals above 90 years, the number of deaths from COVID-19 surpasses the decrease in the number of deaths from pneumonia from 2019 to 2020 in Brazil, by more than twice in all age ranges and even more on average. The same conclusions arise from the graphs in Figure 7. From the graphs in Figure 7 one clearly sees the effects of COVID-19 in the number of deaths, and one also sees that SARI deaths were not reported in 2019 at the extension they are being reported in 2020, and at last, one also observes the reduction in deaths reported from pneumonia in 2020 w.r.t 2019.

**Figure 6.**
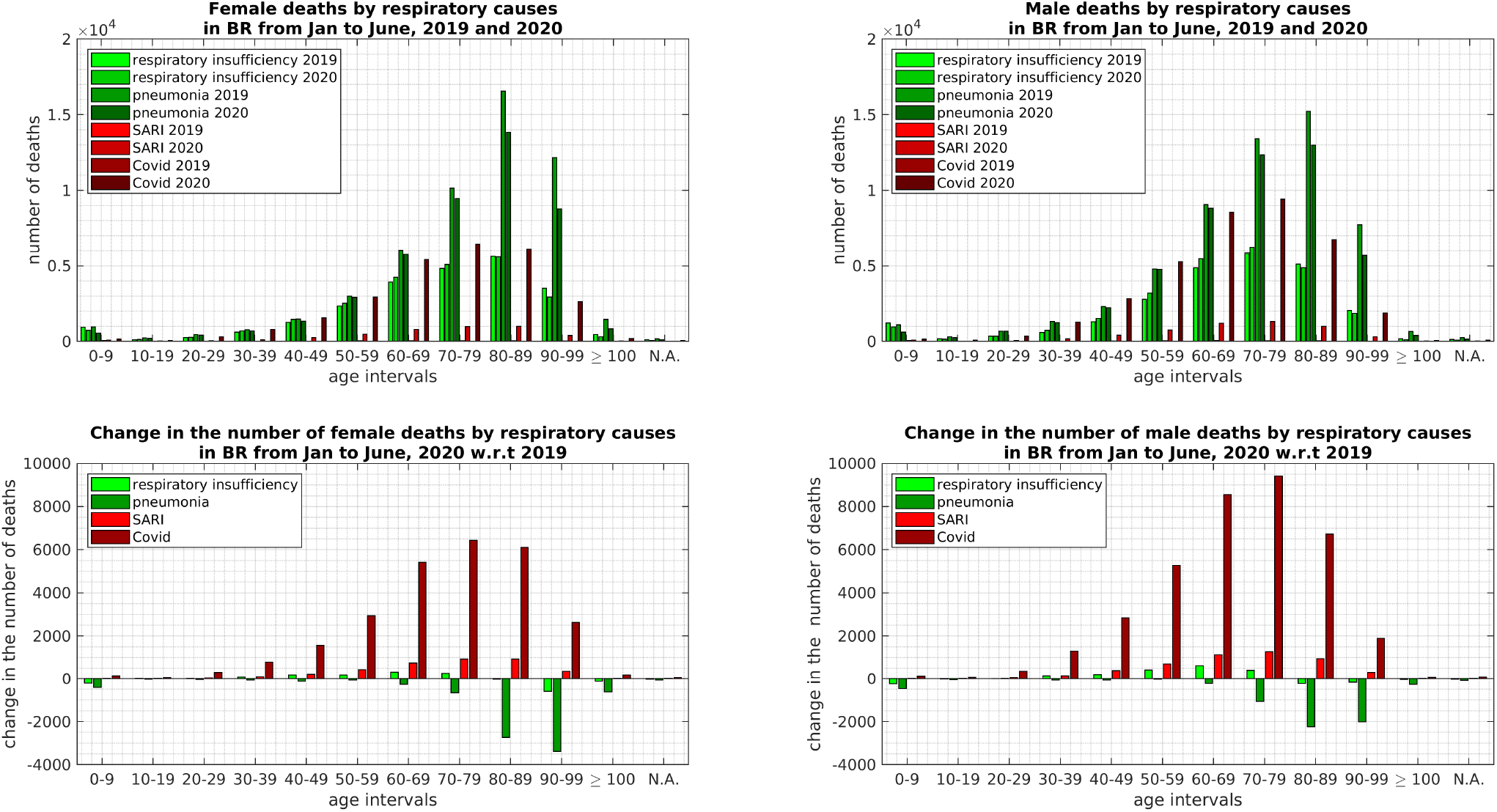
Analysis of deaths from respiratory causes in Brazil, between January and June of 2019 and 2020, for both sexes and different age intervals. At the top left corner, we present the numbers of female deaths resulting from the respiratory causes (respiratory insufficiency, pneumonia, and SARI) and COVID-19. At the top right corner, we present the numbers of male deaths resulting from respiratory causes and COVID-19. At the bottom left corner, we present the changes between 2019 and 2020 in the number of female deaths resulting from respiratory causes and COVID-19. At the bottom right corner, we present the changes between 2019 and 2020 in the number of male deaths resulting from respiratory causes and also COVID-19. Data gathered on July 30th, 2020 from^20^.

**Figure 7.**
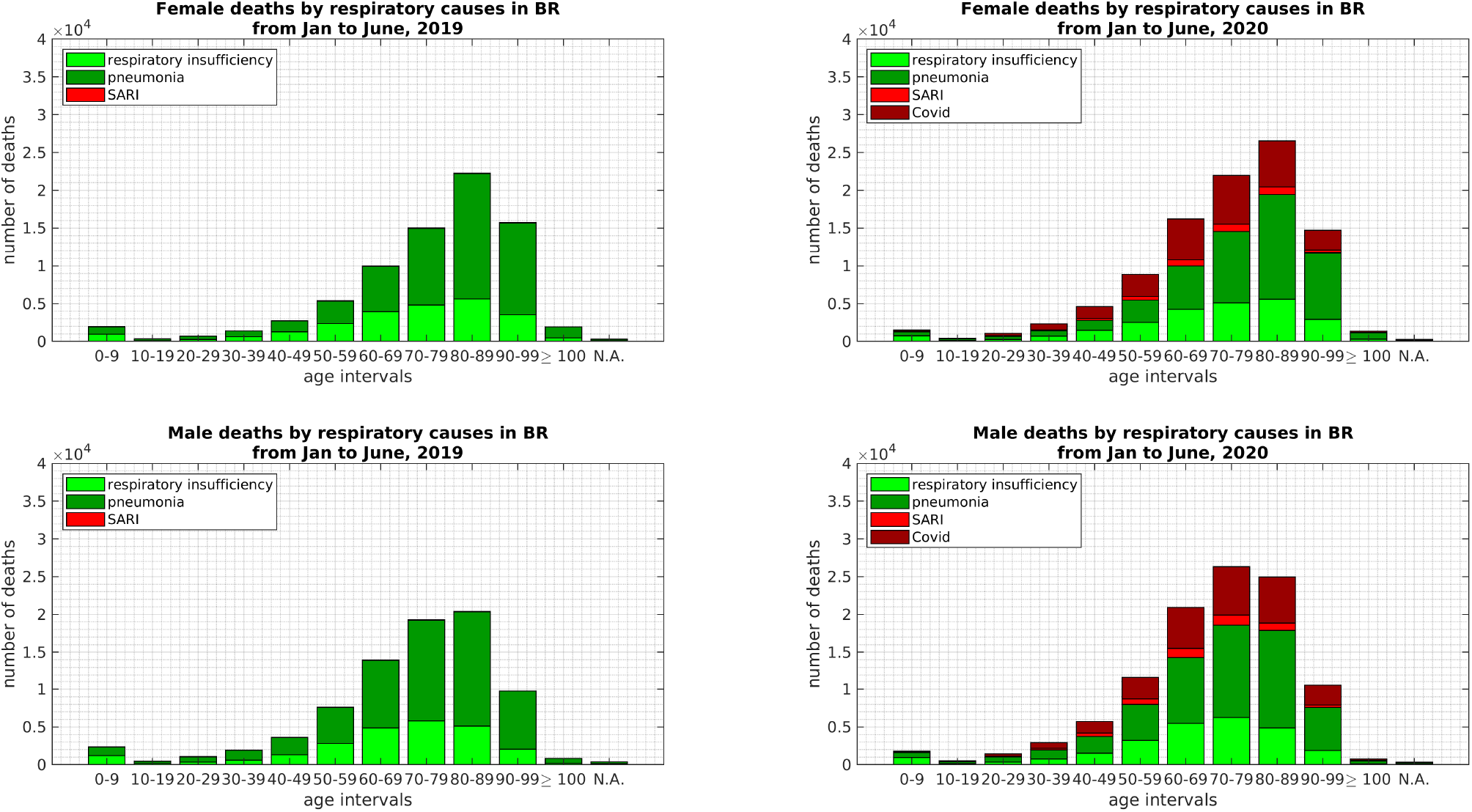
Number of deaths from respiratory causes in Brazil between January and June of 2019 and 2020, for both sexes and ranged by age intervals. At the top left corner, we present the number of female deaths resulting from different respiratory causes (respiratory insufficiency, pneumonia, and SARI) in 2019. At the top right corner, we present the number of female deaths resulting from different respiratory causes in 2020 (including COVID-19). At the bottom left corner, we present the number of male deaths resulting from different respiratory causes in 2019. At the bottom right corner, we present the number of male deaths resulting from different respiratory causes in 2020 (including COVID-19). Data gathered on July 30th, 2020 from^20^.

### Sex influence in death rates

The presented graphs also shows significant differences between sexes in terms of deaths. Studies have shown an association between the patient’s genetic (chromosomal) sex (XX or XY) and mortality rates, as well as the severe form of COVID-19. Overall, the probability of death from COVID-19 is 65 percent higher in males, due to differences in immune responses and inflammatory processes^23,24^.

## Conclusions

The COVID-19 outbreak has increased the death rate in Brazil during April, May, and June in 2020. In some states, this effect is more pronounced, since due to the large area of Brazil, the disease has arrived at different dates in different regions. While in some states the infection is steep, in others it is already declining. Nevertheless, we have observed that the monthly increase in the total number of deaths is largely correlated with the monthly increase in the number of deaths due to respiratory causes. Moreover, the data show that the vast majority of deaths due to respiratory causes are due to COVID-19.

We observed that the increase in deaths due to respiratory causes during the COVID-19 pandemic is more pronounced for ages ranging from 30 to 69 years old, where it lies above 50% for both sexes (considering 10 years intervals). An increase is also observed in the 70-79 and 80-89 age intervals but less pronounced while for the more elder (above 90 years old) fewer deaths were reported in 2020 than in 2019. The young people in the 10-19 and 20-29 years intervals have also died significantly more from respiratory causes in 2020 than in 2019, in the 20-29 age interval, an increase above 40% for both sexes is observed. Altogether they result in a combined percentage increase in the total number of deaths larger than 10% for both sexes, which is larger than the expected increase due to demographics. The data also shows that the increase in the number of deaths is a bit more pronounced for males than females.

In terms of methodology, the present work shows that the analysis of civilian records can be a tool capable of quantitatively revealing peculiarities of the spread of the COVID-19 epidemic in large and heterogeneous countries. It would be interesting to apply the methodology used here in the civil database of other countries with a population of hundreds of millions.

## Data Availability

All data supporting the findings, as well as the computer code used to process and analyze the death data from civil registration, are available at a GitHub Repository.

https://github.com/lisandrolovisolo/deaths_covid19_transparency_BR

## Data and code availability

All data supporting the findings, as well as the computer code used to process and analyze the death data from civil registration, are available at https://github.com/lisandrolovisolo/deaths_covid19_transparency_BR.

## Acknowledgments

The authors thank the financial support from the Brazilian agencies Conselho National de Pesquisa (CNPq), Coordenagao de Aperfeigoamento de Pessoal de Nível Superior - Brasil (CAPES), and the Carlos Chagas Filho Research Foundation of Rio de Janeiro State (FAPERJ). They are also grateful to the *COVID-19: Observatorio Fluminense* team for the collaboration in analyzing other data from the pandemic.

## Author contributions statement

L.L. conceived the research plan. D.C. collected the data. L.L. and D.C. treated the data and generated the graphs. All authors were involved in the analyses and discussions of the results. L.L, A.C., R.B, M.G. and P.C. wrote the manuscript. All authors reviewed and approved the final manuscript.

## Additional information

Competing Interests: The authors declare no competing interests.

